# An acceptable method to evaluate the analytical performance of real-time fluorescent RT-PCR targeting SARS-CoV-2

**DOI:** 10.1101/2020.05.18.20105247

**Authors:** Xiao-Dong Ren, Qi-Mei Tang, Min Chen, Qian Huang, Heng-Liu Huang, Liu Wang, Ning Su, Xian-Ge Sun, Kun Wei, Wei-Ping Lu, Shao-Li Deng, Qing Huang

**Affiliations:** Department of Laboratory Medicine, Daping Hospital, Army Medical University, Chongqing, 400042, P.R.China

**Keywords:** Coronavirus disease 2019, Severe acute respiratory syndrome coronavirus 2, Real-time fluorescent PCR, Analytical performance, Methodology evaluation

## Abstract

It was necessary to carry out methodologies evaluations of real-time fluorescent reverse-transcription PCR (RT-PCR) targeting severe acute respiratory syndrome coronavirus 2 (SARS-CoV-2). Considering biosafety issues and lack of positive specimens in some special locations in China, the routine specimens from healthy individuals were used to perform methodologies evaluations, in which the indexes were the differences of quantification cycle values (Δ *C*_q_) between human derived internal reference control (IRC) genes of a specimen and quality control (QC). Serial experiments were carried out to evaluate various factors that might affect aforementioned methodologies, such as types of virus transport mediums, methods of specimen pretreatment and template preparation, specimen vortex strength, specimen storage temperature and duration. The results showed that using Δ*C*_q_ values as indexes, among various factors that might affect analytical performance, it was better to store specimens in the normal saline transport mediums, inactivate pathogens using water or metal bath, release more virus particles from swabs by vortex mixing, extract nucleic acids with centrifuge methods, and perform amplification assays timely. Aforementioned opinions and optimum conditions were further confirmed by SRAS-CoV-2 pseudovirus and clinical positive specimens. Altogether, the results of this study indicated that the routine specimens from healthy individuals could be used to evaluate the analytical performance of real-time fluorescent RT-PCR targeting SRAS-CoV-2, of which the indexes were the Δ*C*_q_ values between IRC genes of a specimen and QC. This acceptable method was extremely valuable in both theoretical and practical significance under current pandemic of coronavirus disease 2019 (COVID-19).

## Introduction

Since the outbreak of coronavirus disease 2019 (COVID-19) caused by severe acute respiratory syndrome coronavirus 2 (SARS-CoV-2), it has spread rapidly at home and abroad in a short period of time, threatening human health world widely [1]. On March 11, 2020, the World Health Organization announced the global COVID-19 spread with pandemic [2]. Up to May 18, 2020, more than 4.5 million people have been infected in 213 countries, areas and territories around the world, in which about 310,000 individual have died, and the number of infected people continues to rise [3]. Early diagnosis is essential to identify COVID-19 cases in order to provide correct treatment in time and effectively prevent the spread of infectious diseases [4, 5]. In China, from the second edition to the latest seventh edition of Diagnosis and Treatment Program issued by the National Health Committee, all of them pointed out that real-time fluorescent reverse transcription PCR (RT-PCR) is one of the golden standards to confirm COVID-19 suspected patients.

Until now, many scientific research institutions and medical enterprises in China carried out studies to develop SARS-CoV-2 nucleic acid detection kits. Up to May 18, 2020, nearly twenty kinds of kits targeting SARS-CoV-2 were commercially available in China. Because there were various methods to preserve collected oropharyngeal swabs (e.g. various virus transport mediums), pretreat specimens, and prepare nucleic acid templates, it was essential to perform methodology evaluations to screen the optimum conditions to ensure reliable results of commercial kits targeting SARS-CoV-2. However, there were still no studies to suggest how to make aforementioned selections. It might be the reasons that several times of RT-PCR detections were needed to confirm suspected cases of COVID-19 [6, 7]. Therefore, it is extremely necessary to evaluate the relevant methods of SARS-CoV-2 nucleic acid detections, in order to guide the laboratory technicians to select the most appropriate methods to ensure reliable diagnosis results for the clinic.

In standard, SARS-CoV-2 positive specimens should be used to perform methodology evaluations [8, 9]. However, there were only fewer COVID-19 cases outside Hubei province since the outbreak of COVID-19 pandemic in China. It was almost unable to get enough COVID-19 specimens to perform the methodology evaluation for laboratories outside Hubei province. For example, of the approximately 5,000 specimens analyzed in our hospital, only two showed positive results. In general, the amplification of human RNA was used as an internal reference control (IRC) for most commercial kits targeting SARS-CoV-2 [10, 11]. Being infected respiratory epithelial cells, the nucleic acids of SARS-CoV-2 could be co-extracted along with the human-derived IRC templates [12, 13]. Therefore, it was speculated that the amplification efficiencies of IRC genes could be used to reflect indirectly that of targeted SARS-CoV-2.

In this study, the difference of quantification cycle values (Δ*C*_q_) of human derived IRC genes between a specimen and quality control (QC) were used as the indexes to evaluate specific commercial available real-time fluorescent RT-PCR kits targeting SARS-CoV-2. Finally, the optimum conditions were further confirmed with pseudovirus and positive specimens of SARS-CoV-2.

## Materials and Methods

### Collection of various specimens and pseudoviral

All specimens were oropharyngeal swabs (Medico, Guangdong, China), of which negative and positive specimens were come from healthy volunteer and COVID-19 confirmed cases, respectively. This study was approved by the ethics committee of Daping Hospital. Written informed consent was obtained from healthy volunteers, patients or their family members prior to specimen collection. Additionally, pseudovirus carrying N gene fragment of SARS-CoV-2 (Zeesan Biotech, Shanghai, China) was also used in this study. After collections, the specimens were detected in time or stored 4 °C or room temperature (RT) for certain duration.

### Pretreatment of the various specimens and pseudovirus

Three kinds of virus transport mediums were normal saline transport medium (NS; Kelun, Sichuan, China), Nanxin transport medium (NX; Longsee, Guangdong, China), and Youkang transport medium (YK; Yocon, Beijing, China). According to the requirements of various evaluation experiments in followed sections, oropharyngeal swabs collected from healthy volunteers and COVID-19 confirmed cases were stored in at least one of the above mediums. After being vortex mixing for 30s, each specimen was further divided into three parts. Finally, the medium having swabs was inactivated in a water bath at 56 °C for 10 minutes (IWS), one of the other two was inactivated in a metal bath at 56 °C for 10 minutes (IBS), and the last one was not inactivated (NI). Some specimens were spiked with certain concentration of pseudovirus particles.

### Preparation of the nucleic acid templates

After the specimen pretreatment, three kinds of template preparation methods were performed, i.e. magnetic bead (MB), centrifugation (CF) and one-step (OS) method. For MB method, 400 μL of the specimen was used to extract nucleic acids according to manufacturer’s introductions (Sansure Biotech, Hunan, China), and the templates were eluted into 100 μL of sterile water. For CF method, 400 μL of the specimen was centrifuged at 13000 g for 10min followed by adding 100 μL of nucleic acid release agent (Sansure Biotech) to dissolve pellets. For OS method, 50 μL of the specimen was directly and gently mixed with 50 μL of nucleic acid releasing agent (Sansure Biotech).

### Real-time fluorescent RT-PCR

A commercial available SARS-CoV-2 nucleic acid detection kits (Sansure Biotech) were used to analyze various specimens. It could be used to analyze both *ORF1ab* and *N* genes of SARS-CoV-2, as well as the human derived IRC genes (i.e. *Rnse* P). Same QC reactions were included within each batch of experiments to be served as positive or negative controls. All reactions were performed in duplicate to confirm reproducibility. Amplifications were performed on CFX96 PCR System (Bio-Rad; Bio-Rad Lab. Inc., USA) or Cobas Z480 (Roche; Roche Molecular Diagnostics, Pleasanton, CA) instruments. The reaction systems and procedures were carried out according to the instructions of the kits.

### Statistical analysis

The Δ*C*_q_ values of human derived IRC genes between specimen and QC (i.e. Δ*C*_q_ = *C*_qSpedmen_-*C*_qQC_) were used as an index to evaluate amplification efficiencies of the current used commercial RT-PCR kits. Data were analyzed by ANOVA and Tukey’s tests.

## Results

### Analytical performance evaluation of various combinations between specimen pretreatment and template preparation methods

Ten specimens of oropharyngeal swabs collected from healthy individuals were stored in NS mediums. Each specimen was pretreated with three methods (i.e. IWS, IBS, and NI) followed by nucleic acid extractions with three methods (i.e. MB, CF, and OS). All reactions were performed in duplicate on both of Bio-Rad and Roche instruments. A total of about 360 data were generated.

Compared with no difference existed between any specimen pretreatment methods (*p* > 0.05), there were statistical significant differences for Δ*C*_q_ values between various template preparation methods (*p* < 0.05) on both Bio-Rad and Roche instruments (Fig. 1). Moreover, there were no interactions between methods of template preparation and specimen pretreatment (*p* > 0.05; data not shown).

**Figure 1.**
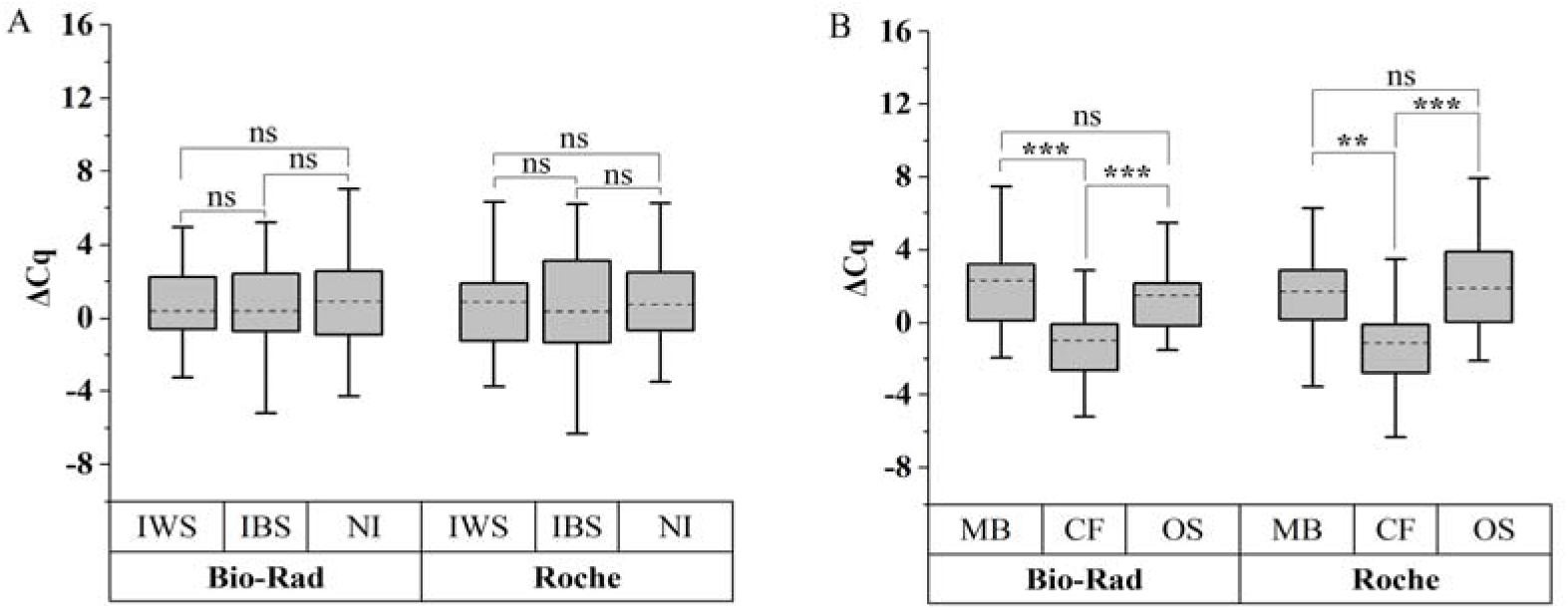
Features of various combined performances between specimen pretreatment and template preparation methods. It showed the IRC Δ*C*_q_ values in box chart plots (n=30) of various specimen pretreatment (panel A) or template preparations (panel B) methods. Parallel amplifications were performed on Bio-Rad and Roche instruments. Asterisks indicate that the *p* value was regarded as a significant difference (i.e. *: *p*<0.05; **: *p*<0.01; ***: *p*<0.001). The ns indicates there is no statistical significant difference.

Further analysis results of various template preparation methods showed that, for both Bio-Rad and Roche instruments, there were statistical significant differences between Δ*C*_q_ values of CF methods and that of both MB and OS methods (*p* < 0.01), and no differences between that of MB and OS methods (*p* > 0.05). The corresponding average Δ*C*_q_ value of CF methods was lower than that of MB and OS methods for both instruments, i.e. 3.209 and 2.576 for Bio-Rad instruments, respectively, and 2.760 and 3.558 for Roche instrument, respectively. These results indicated that CF methods had the best analytical performance under aforementioned various conditions.

### Analytical performance evaluation of various transport mediums and specimen vortex strength

Nine specimens of oropharyngeal swabs were collected from three healthy individuals. Three specimens of each person were stored in NS, YK, and NX virus transport mediums, respectively. And then, for each specimen, various methods of specimen pretreatment and template preparation were performed as mentioned in the previous sections. All reactions were performed in duplicate on both of Bio-Rad and Roche instruments. A total of about 108 data were generated. For various kinds of virus transport mediums (i.e. NS, YK, and NX), the analytical performance of both specimen pretreatment (i.e. IWS, IBS, and NI) and template preparation (i.e. MB, CF, and NI) methods were similar to that of the previous sections. Moreover, there were no interactions between methods of virus transport medium, template preparation and specimen pretreatment (*p* > 0.05).

Further analysis results showed that the transport medium had significant effects on Δ*C*_q_ values for both Bio-Rad and Roche instruments (*p* <0.01). For NS, YK and NX mediums, the average Δ*C*_q_ values of Bio-Rad instruments were 0.801, 1.936 and 3.733, respectively, and that of Roche instruments were 0.777, 1.043 and 2.901, respectively. Statistical analysis results indicated that there were significant differences between Δ*C*_q_ values of various kinds of transport mediums (*p* < 0.05) for both instruments, except for that there were no differences (*p* = 0.865) between that of NS and YK mediums only for Roche instrument (Fig. 2A). Obviously, for both instruments, NS and NX mediums had the best and worst analytical performance, respectively. Notably, for NX mediums, there was no amplification curve in reaction assays using OS methods to prepare templates.

**Figure 2.**
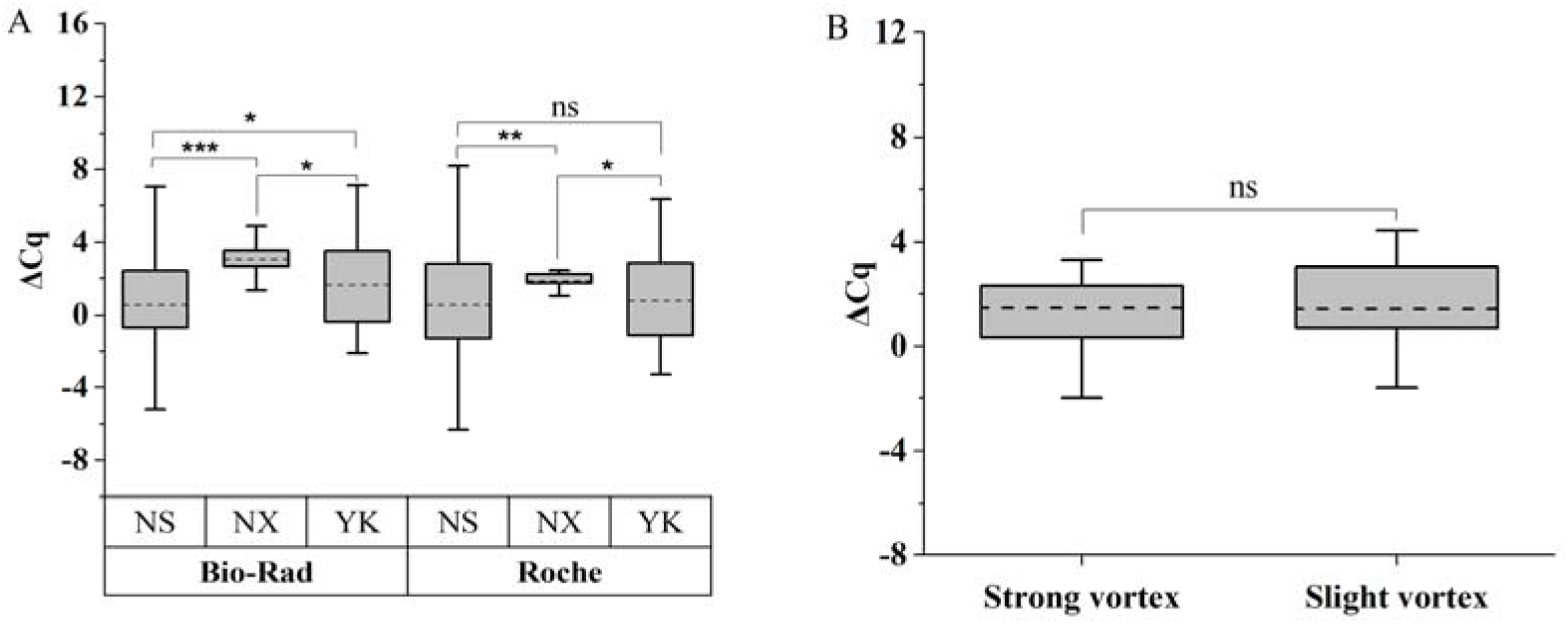
Features of various transport mediums and specimen vortex strength. It showed the IRC Δ*C*_q_ values in box chart plots of various transport mediums (panel A; n=9) or vortex strength (panel B; n=6). Parallel amplifications were performed on Bio-Rad and Roche instruments. Asterisks indicate that the *p* value was regarded as a significant difference (i.e. *: *p*<0.05; **: *p*<0.01; ***: *p*<0.001). The ns indicates there is no statistical significant difference.

In the experiment to evaluate analytical performance of vortex strength, the results showed that the average Δ*C*_q_ value of vigorous mixing was 0.296 smaller than that of gentle mixing. However, there was no statistical difference (*p* = 0.601; Fig. 2B).

### Analytical performance evaluation of storage temperature and duration

Six specimens of oropharyngeal swabs were collected from three healthy individuals. Two specimens of each person were stored in NS and YK virus transport mediums, respectively. And then, each specimen were evenly divided into three parts, of which the first part was tested immediately (i.e. 0h), and the second and third part were stored at 4 °C and RT conditions, respectively, followed by tested at 12h and 24h, respectively. Nucleic acid extractions were carried out using NI specimen pretreatment followed by CF template preparation. All reactions were performed in duplicated on only Bio-Rad instrument. A total of about 60 data were generated.

For both of NS and YK mediums, it was found that the Δ*C*_q_ values were increased with the extension of duration at 4 °C or RT storage temperature. At conditions of 12 or 24 h storage duration, the Δ*C*_q_ values between specimens storage at 4 °C and RT were very close when NS mediums were used (Fig. 3A), and significant different when YK mediums were used (Fig. 3B). These results indicated that the storage temperature had little effects on the analytical performance of NS mediums, and significant effects on that of YK mediums. Based on the comparative analysis of the Δ*C*_q_ values at various storage conditions, it was better to use NS mediums in clinical practice, and storage specimens at 4 °C when YK mediums were used.

**Figure 3.**
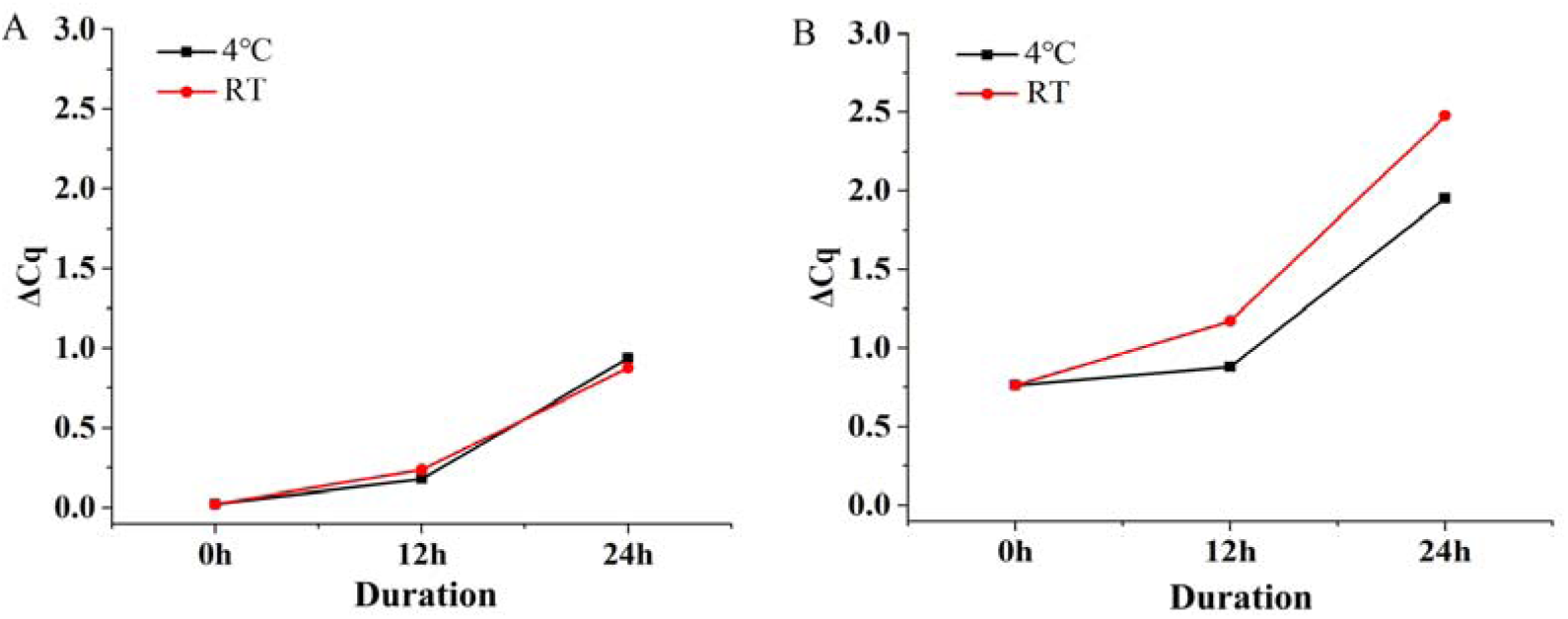
Features of various storage temperature and duration conditions. It showed the average Δ*C*_q_ values (n=3) under conditions of various storage temperatures and durations when NS (panel A) or YK (panel B) mediums were used.

### Clinical evaluation of analytical parameters using pseudovirus and COVID-19 specimens

Two specimens of oropharyngeal swabs from healthy individuals were collected and stored in NS transport mediums. And then, each specimen was evenly divided into two parts. Definite concentrations of pseudovirus carrying fragments of SARS-CoV-2 *N* gene was spiked into pure NS mediums and one part of aforementioned NS mediums containing specimens from healthy individuals. Finally, three types of specimens carrying various templates, i.e. human IRC and SARS-CoV-2 *N* genes (Fig. 4A), purely human IRC genes (Fig. 4B) and SARS-CoV-2 *N* genes (Fig. 4C), were directly used to extract nucleic acids with three certain methods, i.e. CF, MB and OS methods. All reactions were performed in duplicated on only Bio-Rad instrument. A total of about 36 data were generated.

**Figure 4.**
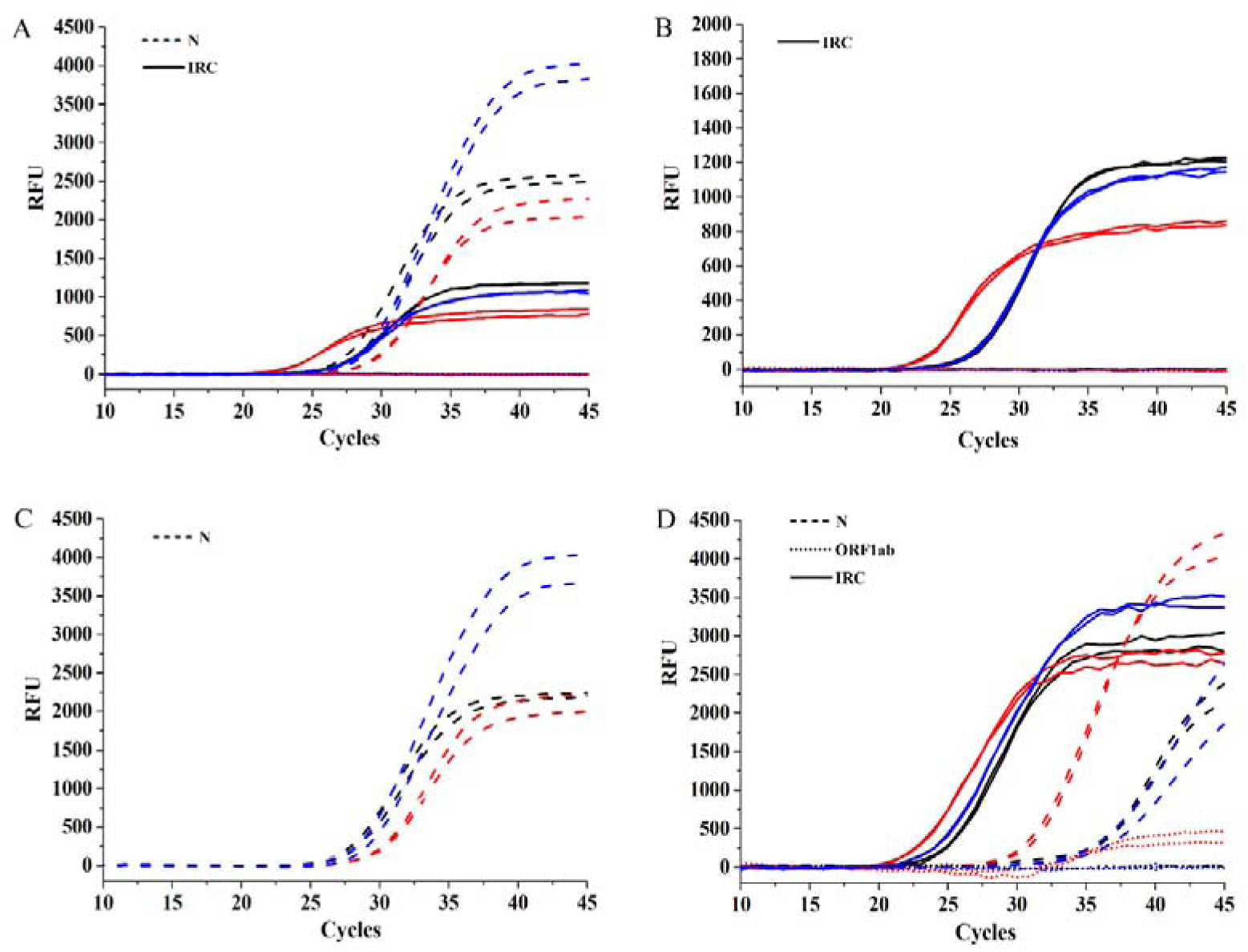
Example results of pseudovirus and SRAS-CoV-2 patient specimens. It showed results of mixture containing various templates, i.e. pseudovirus *N* and human IRC genes (panel A), human IRC genes (panel B), and pseudovirus *N* genes (panel C). The panel D showed results of positive specimens. The methods of template preparation were indicated with various colors, i.e. red, black and blue lines indicated CF, MB and OS methods, respectively.

The results of all specimens showed that there was very close *C_q_* values of the IRC or *N* genes between MB and OS methods (Fig. 4A-C). The *C*_q_ values of the IRC genes of CF methods were smaller than that of both MB and OS methods (Fig. 4A and 4B), which was consistent with previous sections, i.e. CF methods had better analytical performance (Fig. 1 and 2). However, the *C*_q_ values of the *N* genes of CF methods were larger than that of both MB and OS methods (Fig. 4A and 4C), which was inconsistent with that of IRC genes. Moreover, it showed similar results for *N* gene even increased the centrifuge force and time (data not shown). This might be because that the pseudovirus could not be concentrated with human cells together by CF methods.

Finally, two specimens of oropharyngeal swabs from COVID-19 confirmed cases were collected and stored in NS transport mediums. They were directly used to extract nucleic acids with three certain methods, i.e. CF, MB and OS methods. The results of all specimens showed that there was very close *C*_q_ values of the IRC or *N* genes between MB and OS methods (Fig. 4D). However, the *C*_q_ values of the IRC and *N* genes of CF methods were both smaller than that of MB and OS methods (Fig. 4D), which were consistent with analytical performance using human IRC as an index in previous sections. Moreover, it was notably that there were visible amplification curves of SARS-CoV-2 *ORF1ab* genes only in reactions using CF method (Fig. 4D). These results further confirmed that the CF method was better than both MB and OS method, and strong suggested that the amplification efficiencies of IRC genes could be used to evaluate the analytical performance of real-time fluorescent RT-PCR kits targeting SRAS-CoV-2.

## Discussion

Under the current special epidemic conditions, as golden standard to diagnose COVID-19 suspected cases, real-time fluorescent RT-PCR targeting SARS-CoV-2 was routinely implemented in most of the large- and medium-sized laboratories in China [14-16]. There are several types of specimens that could be used to detect SARS-CoV-2 nucleic acids, such as respiratory nasal swabs, oropharyngeal swabs, sputum, and so on [17-19]. Moreover, there were also many factors that might affect the analytical performance of SARS-CoV-2 nucleic acid detection, such as types of virus transport mediums, methods of specimen pretreatment and template preparation, specimen storage temperature and duration. Conventionally, it is better to use SARS-CoV-2 positive specimens to evaluate performance of real-time fluorescent RT-PCR kits [8, 9]. However, considering the biosafety and lack of positive specimens in most laboratories outside Hubei province in China, it was needed to find out alternative methods to evaluate analytical performance of SRAS-CoV-2 nucleic acid detections.

As the virus to infect human respiratory epithelial cells, SARS-CoV-2 particles could be co-extracted with human cells containing the IRC genes that could be detected by most of the commercial available real-time fluorescent RT-PCR kits [10, 11]. Therefore, the abundance of IRC genes could be theoretically used to represent the extraction or preparation efficiency of SARS-CoV-2 nucleic acid. Compared with conventional strategies using specimens from COVID-19 confirmed cases, the current evaluation methods using specimens from healthy individuals had advantages as follows at least: much simpler and safer to collect various specimens, and more easily to perform evaluations just in routine clinical laboratories without considering biosafety issues. Based on above opinions, in this study, the specimens of oropharyngeal swabs collected from healthy individuals were used to evaluate the analytical performance of SARS-CoV-2 nucleic acid detections, in which the indexes were the Δ*C*_q_ values between IRC genes of a specimen and QC. Serial experiments were carried out to evaluate various factors that might affect analytical performance, such as methods of specimen pretreatment and template preparation, types of virus transport medium, storage temperature and durations. Being extremely valuable in both theoretical and practical significance, the analytical performances indicated by aforementioned Δ*C*_q_ values were finally confirmed by pseudovirus and COVID-19 specimens.

Compared with both MB and OS methods of template preparations, the CF methods showed the best analytical performance. Using larger initial volume of specimens, the CF methods exhibited theoretical advantages and practical results than OS methods (Fig. 1-4). Moreover, although using same initial volumes of specimens, the CF methods also showed better performance than MB methods (Fig. 1-4), which might be associated with the relative lower recovery rate of MB methods [19]. Notably, in our clinical practice, the CF methods also exhibited reliable results for one another commercial available real-time fluorescent RT-PCR kit (Daan Gene, Guangdong, China; data not shown).

In the experiments using templates containing pseudovirus fragments, the *C_q_* values of IRC or *N* genes were very close, which further confirmed aforementioned conclusions, i.e. the MB methods had low extraction efficiencies, because larger initial volumes of specimens were used in MB methods. However, compared with MB and OS methods, it was interesting to see that the CF methods exhibited lower amplification efficiencies for *N* genes (Fig. 4A and 4C), which was inconsistent with the results indicated by IRC genes (Fig. 1-3 and 4B). The reasons might be that the pseudovirus particle could not be deposited well in CF methods, and therefore, resulting in poor amplification. However, the above problems could not be existed in clinical specimens. The reasons might be that the SARS-CoV-2 mainly existed in the epithelial cells, or adhered to the cell surface. Therefore, the SARS-CoV-2 particles could be co-deposited with the epithelia cells when CF methods were used [12, 13]. The speculations mentioned above were confirmed by subsequent experiments using clinical COVID-19 specimens, in which both IRC and *N* genes exhibited better amplifications in CF method, and the amplification of *ORF1ab* genes only exhibited in CF methods (Fig. 4D). Aforementioned results suggested that the methodology evaluations based on human IRC genes could be implemented as acceptable strategies to explore analytical performance of SARS-CoV-2 nucleic acid detections.

A common limitation to PCR-based method is failed amplification due to the presence of PCR-inhibitory substances in the specimens, such as heme compounds found in blood, aqueous and vitreous humors, heparin, urine, polyamines, plant polysaccharides [20]. Based on the current comparative analysis on various types of virus transport mediums, it was essential to select the types of virus transport mediums when CF or OS method of template preparation was planned to be used in real-time fluorescent RT-PCR targeting certain pathogens. For example, due to the presence of amplification inhibitor, i.e., guanidine salt [21, 22], the analytical performance of NX mediums was always lower than that of both NS and YK mediums (Fig. 2A).

Considering biosafety of RT-PCR detections targeting SARS-CoV-2, it was better to inactivate pathogens using water or metal bath before any followed procedures because there were no significant differences between various methods of specimen pretreatment (Fig 1). For the other factors that might affect the analytical performance of SARS-CoV-2 nucleic acid detections, it was better to implement analysis in time (Fig. 3), and release SARS-CoV-2 particles from oropharyngeal swabs by vortex mixing (Fig. 2B).

## Conclusion

The routine specimens from healthy individuals could be used to evaluate the analytical performance of real-time fluorescent RT-PCR targeting SRAS-CoV-2, of which the indexes were the Δ*C*_q_ values of IRC genes between specimen and QC. Among various factors that might affect performance mentioned before, it was better to store specimens in NS mediums, inactivate pathogens using water or metal bath, release more virus particles from swabs by vortex mixing, extract nucleic acids with CF methods, and perform amplification assays timely. Being extremely valuable in both theoretical and practical significance, aforementioned opinions and optimum conditions were further confirmed by pseudovirus and clinical positive SRAS-CoV-2 specimens.

## Data Availability

All data generated or analyzed during this study are included in this article.

## Acknowledgements

This study was partly supported by the 2019-nCoV Emergency Research Project (No. CWS20C008), the Military Logistics Scientific Research Project (No. 2019HQZX06), the Military Medical Frontier Innovation Ability Training Program (No. 2019CXJSB005) and University Outstanding Talent Support Program.

## Nonstandard Abbreviations

RT-PCR: real-time fluorescent reverse-transcription PCR
SARS-CoV-2: severe acute respiratory syndrome coronavirus 2
Δ*C*_q_: differences of quantification cycle
IRC: internal reference control
QC: quality control
COVID-19: coronavirus disease 2019
RT: room temperature
NS: normal saline
NX: Longsee transport medium
YK: Yocon transport medium
IWS: inactivation using water bath with swab
IBS: inactivation using metal bath without swab
NI: non-inactivation
MB: magnetic beads method
CF: centrifugation method
OS: one-step method
Rnase P: Ribonuclease P
Bio-Rad: Bio-Rad CFX96 thermocycler
Roche: Roche cobas z480 thermocycler

